# Assessing the role of rare pathogenic variants in heart failure progression by exome sequencing in 8,089 patients

**DOI:** 10.1101/2023.07.28.23293350

**Authors:** Olympe Chazara, Marie-Pierre Dubé, Quanli Wang, Lawrence Middleton, Dimitrios Vitsios, Anna Walentinsson, Qing-Dong Wang, Kenny M. Hansson, Christopher B. Granger, John Kjekshus, Carolina Haefliger, Jean-Claude Tardif, Dirk S. Paul, Keren Carss

**Author notes:** Correspondence: Dirk S. Paul. These authors contributed equally (Dirk S. Paul, Keren Carss).

## Abstract

Most therapeutic development is targeted at slowing disease progression, often long after the initiating events of disease incidence. Heart failure is a chronic, life-threatening disease and the most common reason for hospital admission in people over 65 years of age. Genetic factors that influence heart failure progression have not yet been identified. We performed an exome-wide association study in 8,089 patients with heart failure across two clinical trials, CHARM and CORONA, and one population-based cohort, the UK Biobank. We assessed the genetic determinants of the outcomes ‘time to cardiovascular death’ and ‘time to cardiovascular death and/or hospitalisation’, identifying seven independent exome-wide-significant associated genes, *FAM221A*, *CUTC*, *IFIT5*, *STIMATE*, *TAS2R20*, *CALB2* and *BLK*. Leveraging public genomic data resources, transcriptomic and pathway analyses, as well as a machine-learning approach, we annotated and prioritised the identified genes for further target validation experiments. Together, these findings advance our understanding of the molecular underpinnings of heart failure progression and reveal putative new candidate therapeutic targets.

## Introduction

Heart failure is a common, chronic health condition with increasing prevalence. In 2017, an estimated 64.3 million people worldwide were living with heart failure^1^. Around 2% of adults have heart failure and, in those over the age of 65, the prevalence increases to 6–10%^2–5^. Although survival after diagnosis has improved, death rates remain high, with 5-year survival averaging 50%^6^. Heart failure is the leading cause of both hospitalization and readmission amongst older adults^7–9^.

Studies have estimated the heritability of heart failure at 26%^10^. Genome-wide association studies (GWAS) have characterised the contribution of common genetic variants to the aetiology of heart failure^11–13^. We recently described the distinct contribution of rare genetic variation to all-cause heart failure in a case-control study^14^. We also investigate the genetic differences between heart failure subtypes based on left ventricular ejection fraction (LVEF) and the burden of Mendelian cardiomyopathy variants in these patients. We observed an enrichment of variants typically associated with dilated cardiomyopathy in patients with ischemic heart failure, in particular, protein-truncating variants in the *TTN* gene. Our data supported the notion that genes linked to Mendelian cardiomyopathy could represent therapeutic targets for a broader heart failure indication. Collectively, the vast majority of genetic studies for common chronic diseases such as heart failure have informed on disease incidence, with an unclear relationship to disease progression.

Most therapeutic development is targeted at slowing or arresting disease progression in individuals who already have disease. However, studies of disease progression remain challenging, as they require extensive, longitudinal clinical data that require expert curation, such as diagnoses, hospitalisation dates, causes of hospitalisation and death. They also present statistical and computational challenges, as the most well-known approach for analysis of survival (or time-to-event) analysis, the Cox proportional hazards model^15^, does not scale well to large datasets^16^. Partially overcoming these challenges, powerful and efficient time-to-event analysis frameworks have recently been applied to the UK Biobank^17–19^. For heart failure, these studies have mostly focused on time-to-diagnosis phenotypes and/or considered only common genetic variants^20^.

The aim of this study was to identify rare variants associated with heart failure progression. Rare variants have typically larger effects on phenotypes and reveal more direct insights into biology that can be exploited for medicine development^21^. Specifically, the objectives of this study were to (1) perform a survival analysis of heart failure patients to identify genes with an excess or depletion of rare variants in individuals with a detrimental outcome such as shorter time to cardiovascular death; (2) explore how these results can inform the genetic architecture of heart failure progression; and (3) evaluate which of the identified progression-associated candidate genes could present potential candidates for therapeutic development. To address these objectives, we analysed whole-exome sequencing data from patients with heart failure, representing all broad clinical subtypes, from the Candesartan in Heart Failure-Assessment of Reduction in Mortality and Morbidity (CHARM) and Controlled Rosuvastatin Multinational Trial in Heart Failure (CORONA) clinical trials, and heart failure patients from the UK Biobank^22–24^. All three cohort studies had extensive clinical information available and had been previously studied for heart failure incidence. We applied gene-based collapsing analyses within each study, followed by meta-analysis to discover genes with an excess of rare variants associated with two heart failure outcomes, ‘time to cardiovascular death’ and ‘time to cardiovascular death and/or hospitalisation’.

## Results

### Description of the study cohorts and analytical approach

In this study, we analysed a total of 8,089 heart failure patients: 2,672 from CHARM, 2,776 from CORONA and 2,641 from the UK Biobank. From these 8,089 individuals, 1,328 (16.4%) died of cardiovascular death, and 2,451 (30.3%) either died of cardiovascular death or were hospitalised due to heart failure following study entry (**Table 1**, **Figure 1**).

**Figure 1.**
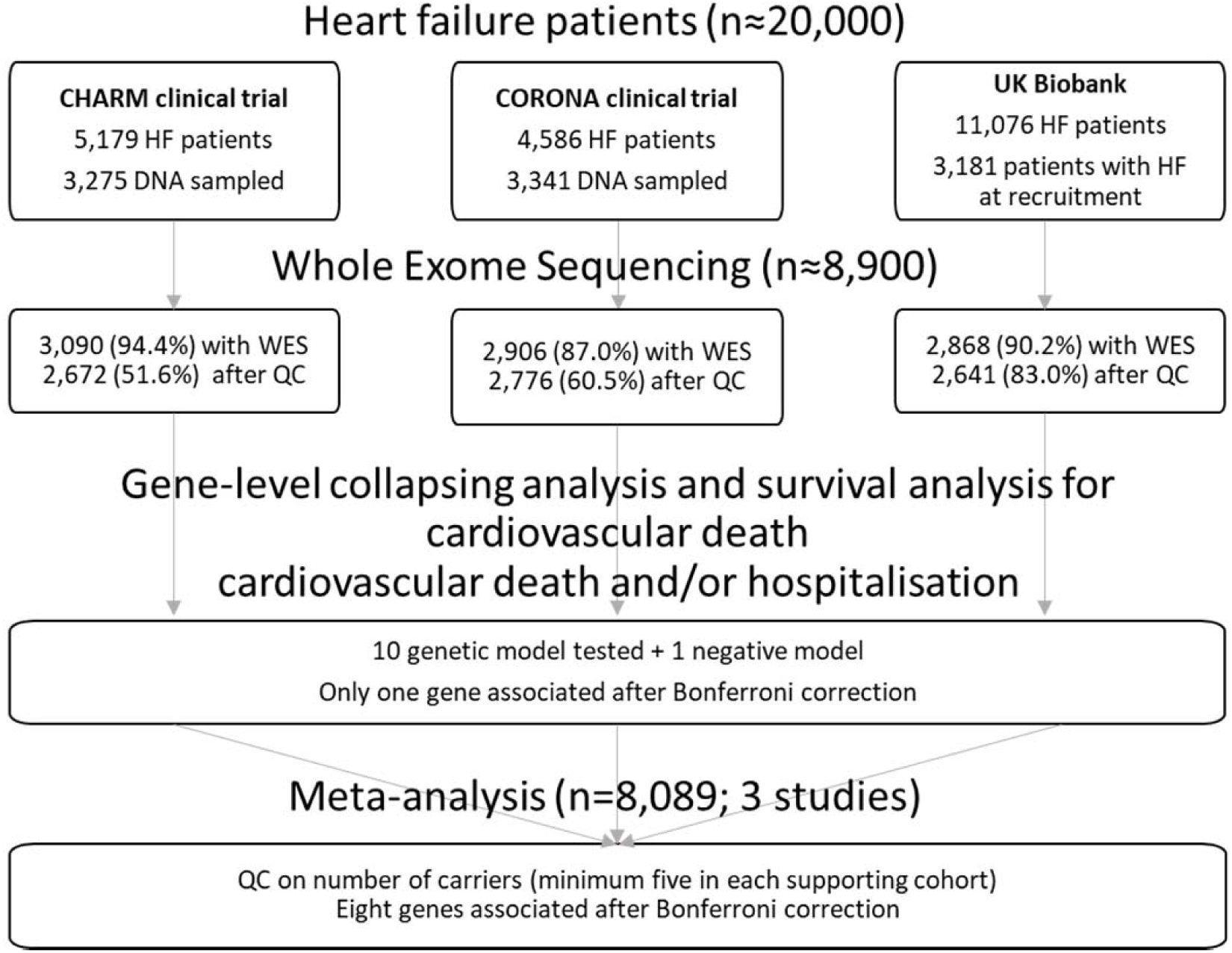
Study design.

**Table 1.**
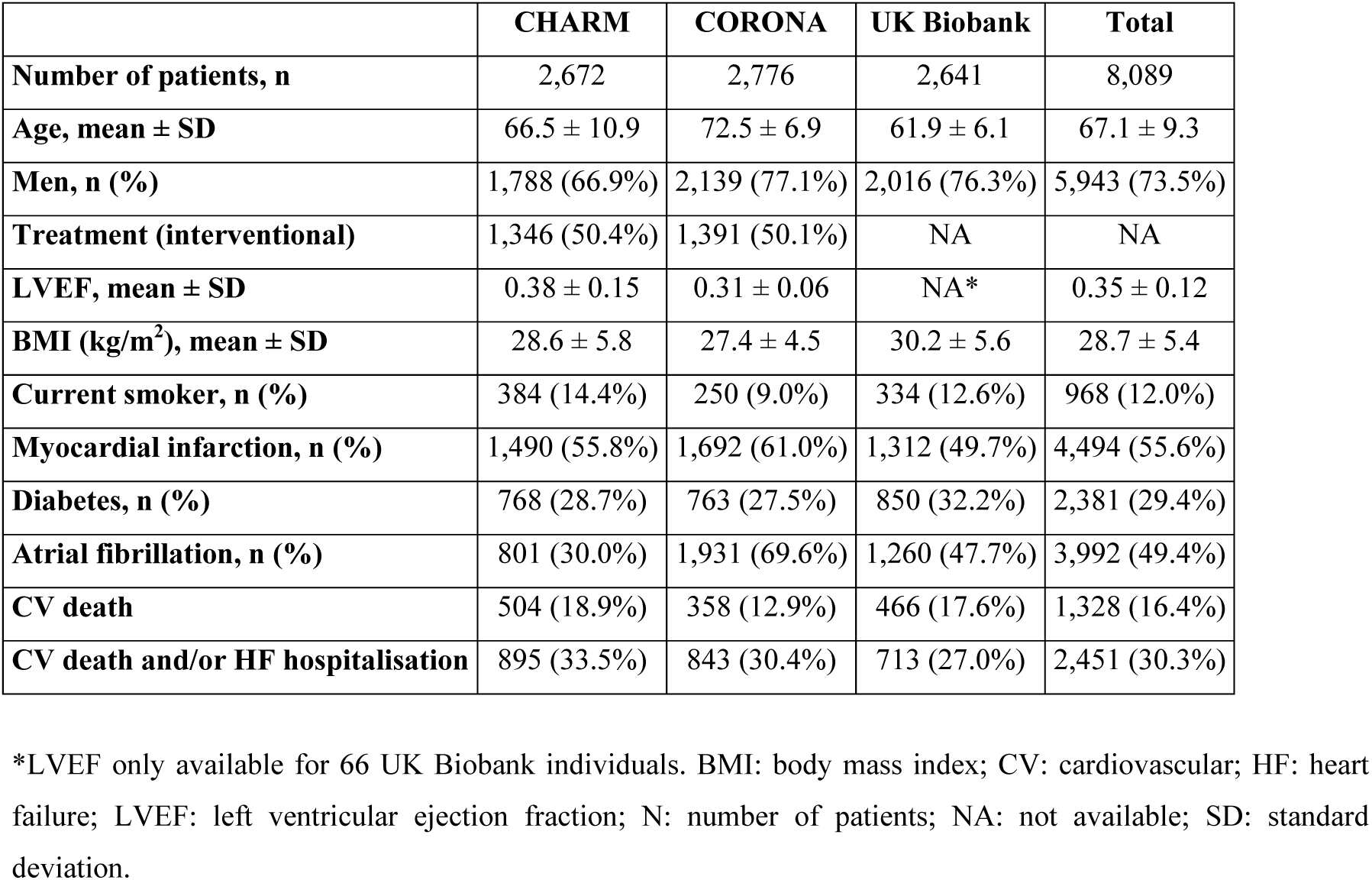
Characteristics of patients included in the analysis.

First, we analysed the three study cohorts separately to detect rare variants associated with the two outcomes ‘time to cardiovascular death’ and ‘time to cardiovascular death and/or heart failure hospitalisation’. We used gene-level collapsing analyses to classify the genotypes, and Cox proportional hazards regression with Firth’s penalized likelihood to analyse each individual’s disease progression (**Methods**). In gene-level collapsing analyses, the proportion of cases with a qualifying variant is compared with the proportion of controls with a qualifying variant in each gene. We applied ten different sets of qualifying variant filters (models) (**Methods**). Finally, we used a meta-analysis approach to identify genes with overall evidence of association.

### Rare-variant analysis for time to cardiovascular death in heart failure patients

For time to cardiovascular death in heart failure patients, the gene-based collapsing meta-analysis identified five significantly associated genes (p<2×10^-7^): *FAM221A* (p=3.85×10^-8^, hazard ratio (HR) 5.14), *CUTC* (p=3.90×10^-8^, HR 5.13), *IFIT5* (p=4.77×10^-8^, HR 4.77), *STIMATE* (p=7.68×10^-8^, HR 4.39) and *TAS2R20* (p=2.57×10^-7^, HR 3.93) (**Table 2**, **Figure 2A-E**). The meta-analysis for these five genes was based on the survival analysis of the three contributing studies, with at least five carriers per study. None of the single-study analysis results for time to cardiovascular death in heart failure patients passed the significance threshold of p<2×10^-7^, demonstrating the power of meta-analysis for this phenotype to discover novel rare-variant based signals (**Supplementary Tables 1-3**). The meta-analysis also revealed a significant association for *CHRNA9* (p=1.89×10^-7^, HR 4.72) but the supporting survival analysis in CHARM was violating the proportionality hazard assumption; therefore, we did not report this association result.

**Figure 2.**
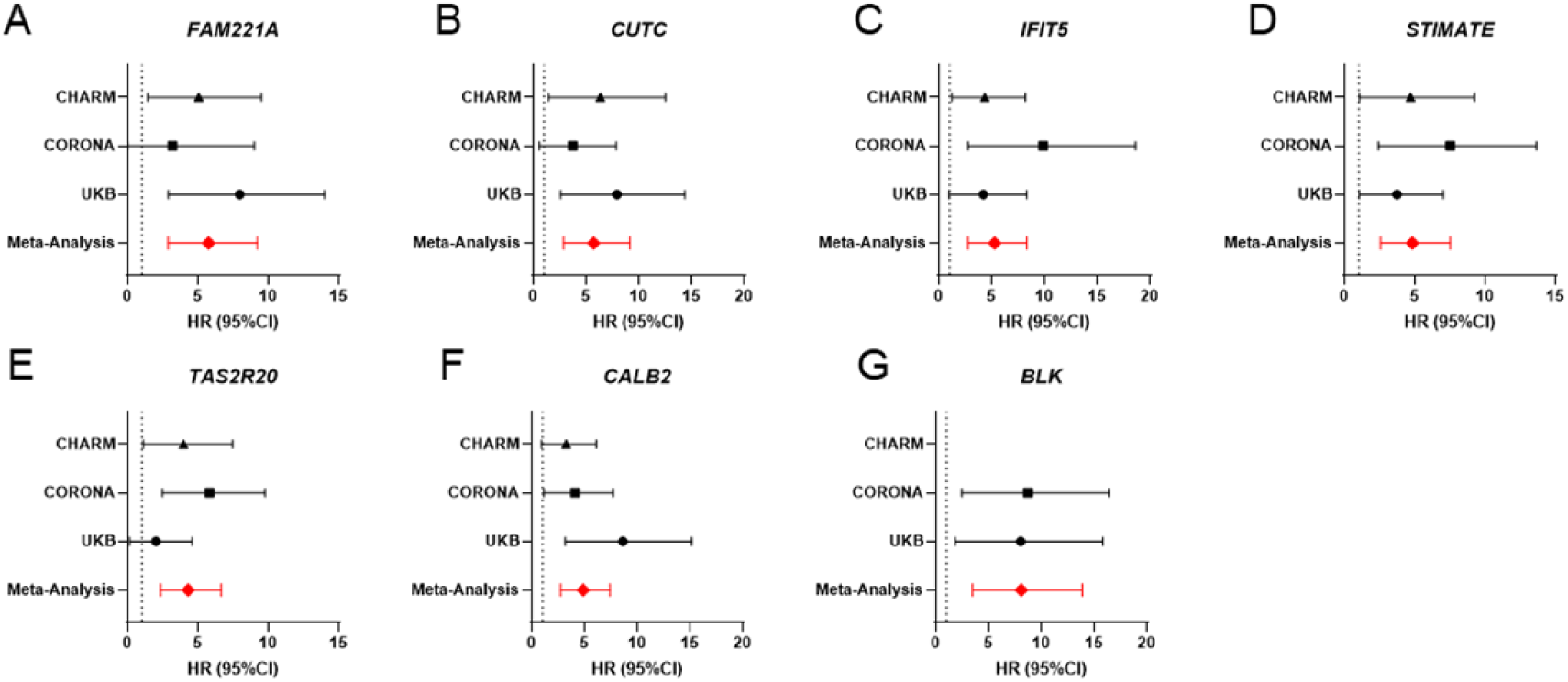
Forest plot of the significant results (p<2.7×10^-7^) from the meta-analysis of CHARM, CORONA and the UK Biobank for time to cardiovascular death (A-E) and time to cardiovascular death and/or hospitalisation for heart failure (F-G). **A**, *FAM221A* in the flexible non-synonymous model; **B**, *CUTC* in the flexible non-synonymous (MTR) model; **C**, *IFIT5* in the flexible damaging model; **D**, *STIMATE* in the flexible non-synonymous model; **E**, *TAS2R20* in the flexible non-synonymous and/or in flexible non-synonymous (MTR) model; **F**, *CALB2* in the flexible non-synonymous model; **G**, *BLK* in the ultra-rare model.

**Table 2.** Gene-based collapsing meta-analysis results for ‘time to cardiovascular death’ (top) or ‘time to cardiovascular death and/or hospitalisation for heart failure’ (bottom).

| Gene | Genetic model* | N (studies) | P | HR (95% CI) | SE | Q | I<sup>2</sup>** | Total<br>N (carriers) | CHARM<br>P (carriers)<sup>†</sup> | CORONA<br>P (carriers)<sup>†</sup> | UK Biobank<br>P (carriers)<sup>†</sup> |
| --- | --- | --- | --- | --- | --- | --- | --- | --- | --- | --- | --- |
| **Time to cardiovascular death** |  |  |  |  |  |  |  |  |  |  |  |
| *FAM221A* | Flexible non-synonymous | 3 | 3.85E-08 | 5.14 (2.87, 9.21) | 1.35 | 0.20 | 38.02 | 25 | 1.38E-02 (8) | 6.48E-01 (7) | 1.77E-04 (10) |
| *CUTC* | Flexible non-synonymous (MTR) | 3 | 3.90E-08 | 5.13 (2.85, 9.13) | 1.35 | 0.50 | 0 | 19 | 1.67E-02 (5) | 1.70E-01 (6) | 6.32E-04 (8) |
| *IFIT5* | Flexible damaging | 3 | 4.77E-08 | 4.77 (2.72, 8.33) | 1.34 | 0.36 | 2.20 | 24 | 2.47E-02 (8) | 9.19E-04 (8) | 5.91E-02 (8) |
| *STIMATE* | Flexible non-synonymous | 3 | 7.68E-08 | 4.39 (2.56, 7.52) | 1.32 | 0.50 | 0 | 25 | 4.28E-02 (6) | 8.69E-04 (10) | 4.28E-02 (9) |
| *TAS2R20* | Flexible non-synonymous,<br>Flexible non-synonymous (MTR) | 3 | 2.57E-07 | 3.93 (2.34, 6.63) | 1.31 | 0.26 | 25.85 | 34 | 3.44E-02 (10) | 1.54E-04 (16) | 7.69E-01 (8) |
| **Time to cardiovascular death and/or hospitalisation for heart failure** |  |  |  |  |  |  |  |  |  |  |  |
| *CALB2* | Flexible non-synonymous | 3 | 4.82E-09 | 4.47 (2.71, 7.39) | 1.30 | 0.20 | 37.83 | 21 | 6.94E-02 (8) | 3.05E-02 (6) | 1.07E-04 (7) |
| *BLK* | Ultra-rare | 2 | 5.46E-08 | 6.88 (3.43, 13.84) | 1.43 | 0.86 | 0 | 10 | NA | 1.49E-03 (5) | 7.67E-03 (5) |

We assessed the distributions of the qualifying variants along the length of each of the five identified genes (**Figure 3A-E**). We found that the observed associations with the lowest p-values were driven by at least eight variants per gene, and were evenly distributed across the length of the gene sequence (**Supplementary Table 4**).

**Figure 3.**
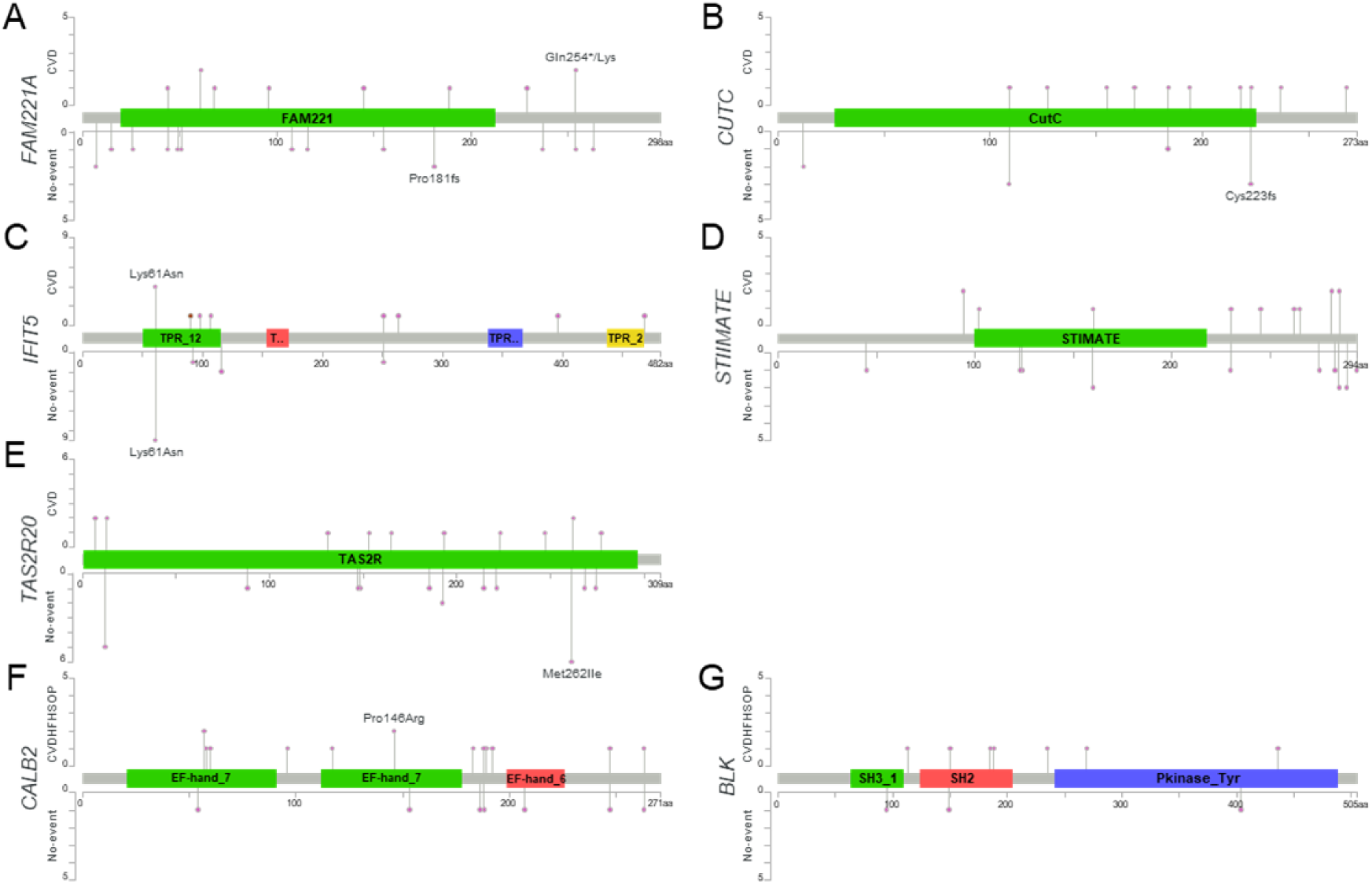
Lollipop plots depicting the individual variants across the candidate genes from the meta-analysis of time to cardiovascular death (A-E) and time to cardiovascular death and/or hospitalisation for heart failure (F-G). Variants in patients with event at the top and no-event at the bottom. More details on variants identified in heart failure patients with detrimental events in **Supplementary Table 4**. **A**, *FAM221A* in the flexible non-synonymous model (ENST00000344962); **B**, *CUTC* in the flexible non-synonymous (MTR) model (ENST00000370476); **C**, *IFIT5* in the flexible damaging model (ENST00000371795); **D**, *STIMATE* in the flexible non-synonymous model (ENST00000355083); **E**, *TAS2R20* in the flexible non-synonymous and/or in flexible non-synonymous (MTR) model (ENST00000538986); **F**, *CALB2* in the flexible non-synonymous model (ENST00000302628); **G**, *BLK* in the ultra-rare model (ENST00000259089). Lollipop plots were produced with the cbioportal mutation mapper tool^61, 62^.

### Rare-variant analysis for the composite of time to cardiovascular death and/or heart failure hospitalisation

For the second of the two outcomes investigates, the composite of time to cardiovascular death and/or heart failure hospitalisation, the gene-based collapsing meta-analysis identified two significantly associated genes (p<2×10^-7^): *CALB2* (p=4.82×10^-9^, HR 4.47) and *BLK* (p=5.46×10^-8^, HR 6.91) (**Table 2**, **Figure 2F-G**). While the meta-analysis for *CALB2* result was based on the survival analysis of the three studies, with at least five carriers per study, the result for *BLK* was only based on the CORONA and UK Biobank studies, as there was only one carrier for *BLK* in CHARM. The proportionality hazard assumption was not met for *CALB2* in CHARM, but after removal of the study from the analysis, the meta-analysis result remained significant (p=2.15×10^-8^, HR 5.48).

We report one significant result from the single-study analysis for time to cardiovascular death and/or heart failure hospitalisation in heart failure patients: *RAD54L* in the flexible non-synonymous model in CHARM (p=6.64×10^-8^, HR 3.96; **Supplementary Tables 5-7**). *RAD54L* in the corresponding analysis in CORONA and the UK Biobank was not significantly associated with heart failure progression (p>0.1 and number of qualifying variant carriers >30 in both cohorts). Different genetic architecture across the studies could reflect underlying clinical differences, due to the differing recruitment criteria employed.

We assessed the distributions of the qualifying variants along the length of the identified genes (**Figure 3F-G**) and found that the observed associations with the lowest p-values were driven by 13 variants for the *CALB2* gene and seven variants for *BLK* (**Supplementary Table 4**). We found these variants to be evenly distributed across the length of the gene sequence (**Supplementary Table 4**).

### Orthogonal genetic evidence in support of rare-variant meta-analysis results

To investigate to what extent the discovered genes from our rare-variant meta-analyses are associated with related phenotypes in complementary data sources, we annotated the genes with orthogonal genetic evidence: (1) curated common-variant association studies for cardiovascular diseases from the Cardiovascular Disease Knowledge Portal (https://cvd.hugeamp.org/); (2) known Mendelian cardiomyopathy-causing genes; and (3) a gene prioritisation method that leverages external data sources using a machine-learning (ML) approach (Mantis-ML)^25^.

We collated the curated evidence from published GWAS from the Cardiovascular Disease Knowledge Portal for the seven candidate genes from the meta-analysis for both tested outcomes (**Table 3**). We found genome-wide significant associations with cardiovascular traits in the genomic regions of *CUTC*, *IFIT5*, *STIMATE*, *CALB2* and *BLK*. There were also genome-wide significant associations with lipids traits in the regions of *CUTC*, *STIMATE*, *CALB2* and *BLK*, and with anthropometric traits in the regions of *FAM211A*, *CUTC*, *STIMATE*, *CALB2* and *BLK* (**Table 3A**). A limitation of this comparison is that a majority of the reported associations had the UK Biobank included in the meta-analysis, and the relationship between the GWAS index variants and causal genes at the loci requires further experimental confirmation. However, we did not detect genome-wide significant associations in either the region or the gene for the most relevant reported phenotypes, i.e., heart failure, non-ischemic cardiomyopathy, atrial fibrillation, any cardiovascular disease, myocardial infarction, and hypertension. Only *IFIT5*, *STIMATE* and *CALB2* had suggestive associations (p<1×10^-5^) for any cardiovascular disease (*IFIT5* and *STIMATE*) or hypertension (*STIMATE* and *CALB2*) (**Table 3B**). The lowest p-values observed in the *CUTC* region were for heart rate (p=2.73×10^-9^), height (p=1.80×10^-28^) and atrial fibrillation (p=1.79×10^-4^) (**Table 3B**).

**Table 3.** Summary of supporting genetic evidence from (A) Cardiovascular Disease Knowledge Portal and (B) published GWAS on several cardiovascular outcomes.

|  | A) Cardiovascular Disease Knowledge Portal |  |  |  |  |  |  |  |  | B) Published GWAS data |  |  |  |  |  |  |  |  |  |  |  |
| --- | --- | --- | --- | --- | --- | --- | --- | --- | --- | --- | --- | --- | --- | --- | --- | --- | --- | --- | --- | --- | --- |
| Gene | Cardiovascular |  |  | Lipids |  |  | Anthropometric |  |  | Heart failure |  | Non-ischemic cardiomyopathy |  | Atrial fibrillation |  | Any cardiovascular disease |  | Myocardial infarction |  | Hypertension |  |
|  | Trait | P | beta | Trait | P | beta | Trait | P | beta | P | beta | P | beta | P | beta | P | beta | P | beta | P | beta |
| *FAM221A* |  |  |  |  |  |  | Height | 3.52E-76 | ▼ -0.0007 | 1.29E-04 | ▲ 1.06 | 5.03E-02 | ▲ 1.07 | 8.76E-05 | ▲ 5.82 | 1.71E-04 | ▲ 1.04 | 9.32E-03 | ▲ 3.38 | 4.57E-03 | ▲ 1.02 |
| *CUTC* | Heart rate | 2.73E-09 | ▼ -0.100 |  |  |  | Height | 1.80E-28 | ▼ -0.0003 | 1.67E-03 | ▲ 1.86 | 8.07E-03 | ▼ 0.85 | 1.79E-04 | ▲ 1.52 | 6.82E-03 | ▼ 0.95 | 1.25E-02 | ▲ 13.52 | 2.20E-05 | ▼ 0.95 |
| *IFIT5* | Diastolic blood pressure | 1.74E-08 | ▲ 0.011 |  |  |  |  |  |  | 2.72E-03 | ▲ 2.59 | 2.37E-03 | ▼ 0.81 | 1.76E-03 | ▲ 1.20 | 2.32E-06 | ▲ 1.02 | 1.62E-02 | ▼ 0.08 | 1.38E-04 | ▲ 1.23 |
| *STIMATE* | Pulse pressure | 1.03E-13 | ▲ 0.007 | TGs | 8.92E-19 | ▼ -0.023 | Height | 8.09E-83 | ▼ -0.0006 | 2.08E-03 | ▲ 1.04 | 1.27E-04 | ▲ 1.20 | 5.00E-03 | ▲ 2.16 | 1.00E-07 | ▼ 0.95 | 7.35E-04 | ▲ 1.17 | 6.26E-06 | ▲ 1.02 |
| *TAS2R20* |  |  |  |  |  |  |  |  |  | 2.21E-03 | ▲ 1.07 | 1.77E-03 | ▲ 1.50 | 4.28E-03 | ▲ 1.73 | 6.80E-03 | ▼ 0.99 | 2.74E-02 | ▼ 0.86 | 4.19E-03 | ▼ 0.86 |
| *CALB2* | Ascending aorta diameter | 2.90E-34 | ▼ -0.190 | LDL-C | 2.57E-292 | ▲ 0.033 | BMI | 2.42E-22 | ▼ -0.020 | 1.34E-03 | ▲ 1.04 | 2.22E-03 | ▼ 0.84 | 8.40E-04 | ▲ 1.37 | 1.22E-04 | ▼ 0.96 | 3.81E-02 | ▼ 0.85 | 3.20E-06 | ▼ 0.96 |
| *BLK* | Systolic blood pressure | 2.07E-22 | ▼ -0.013 | TGs | 9.71E-30 | ▲ 0.027 | BMI | 1.20E-25 | ▼ -0.012 | 7.22E-04 | ▼ 0.94 | 2.72E-03 | ▼ 0.85 | 6.31E-03 | ▲ 1.29 | 1.84E-03 | ▲ 1.02 | 1.27E-03 | ▲ 48.34 | 6.14E-05 | ▲ 1.05 |

From the Cardiovascular Disease Knowledge Portal, we retrieved the most significant common-variant associations ( $p \leq 5 \times 10^{-8}$ ) for cardiovascular, lipid and anthropometric traits at each gene region, clumped by linkage disequilibrium.

Among the known rare or low-frequency variants in cardiomyopathy-causing genes previously identified such as *TTN*, *MYH7* and *MYBPC3* for heart failure incidence and *LMNA* and *LAMP2* for heart failure progression, we observed a non-significant association of ultra-rare variants in *MYBPC3* with time to cardiovascular death (p=2.05×10^-6^, HR 4.38), but not for the other reported genes. Most of these cardiomyopathy-causing genes had a very low number of pathogenic variant carriers, making interpretation challenging. However, *TTN* pathogenic variants were more frequent, indicating that *TTN* might be specifically associated with heart failure incidence rather than progression, which has been reported previously^26^.

Finally, we applied Mantis-ml, a ML framework leveraging publicly available gene annotation data to prioritise disease-gene associations^25^. We used this orthogonal method to investigate the extent to which the significant genes from our rare-variant meta-analyses (p<0.01) are similar to genes known, or predicted with high confidence, to be associated with relevant phenotypes (i.e., the top-ranked genes from Mantis-ml conditioned on heart failure or cardiomyopathy). These similarities were quantified using a step-wise hypergeometric test (**Methods**). For both outcomes, we found the overlap in relevant phenotypes to be greater for the qualifying variant models that capture functional variants than for the synonymous negative control model (**Supplementary Figs. 1A, 1B**). In addition, collapsing the 33 heart failure/cardiomyopathy disease classes available to a single metric (i.e., taking either the maximum or the AUC), we found significantly superior performance of the pooled qualifying variant model compared to the synonymous model for both, time to cardiovascular death (p_max_=1.6×10^-5^ and p_AUC_=8.0 ×10^-4^) and time to cardiovascular death and/or hospitalisation for heart failure (p_max_=5.1×10^-5^ and p_AUC_=3.9×10^-4^) (**Supplementary Fig. 1C**). Taken together, these results showed that the features of the most significantly associated genes from our meta-analysis share similarities with the features of genes already known, or strongly predicted, to be associated with relevant phenotypes, supporting our genetic data.

### Expression analysis in heart tissues aids gene prioritisation for further validation

Next, we assessed the expression patterns of the candidate causal genes underlying the genetic associations for heart failure progression. Normalized gene expression data collected by the GTEx Project^27^ were interrogated for the seven candidates genes from the meta-analysis for both tested outcomes (**Table 2**). We used the 54 different tissues sampled by GTEx as well as a subset of 16 cardiovascular tissues.

The overall tissue expression analysis revealed that *CUTC*, *STIMATE* and *IFIT5* are expressed in almost all tissues analysed; *CALB2* and *BLK* are expressed in multiple tissues but not all; and *TAS2R20* and *FAM221A* have comparatively low expression across all tissues analysed (**Supplementary Fig. 2A**). *CUTC* was the gene most strongly expressed in the heart. In cardiovascular tissue, a similar gene grouping was observed, with *CUTC* highest in skeletal muscle, *CALB2* in adipose, and *BLK* in whole blood and spleen tissues (**Supplementary Fig. 2B**).

Next, we analysed differential expression of the candidate genes in tissues from ventricular myocardial biopsies in heart failure with preserved/reduced ejection fraction (HFpEF/HFrEF), (idiopathic) dilated cardiomyopathy (DCM), hypertrophic cardiomyopathy (HCM), ischemic cardiomyopathy (ICM), and controls from three separate public transcriptomics studies (**Figure 4**, **Supplementary Table 8**)^28^. *CUTC*, *IFIT5*, *TAS2R20*, *FAM221A* and *BLK* showed significant differential expression between the healthy and diseased heart. Among these, *CUTC* was most consistently dysregulated, being downregulated across studies and heart failure aetiologies. *TAS2R20* and *BLK* have low expression in the heart, making the results challenging to interpret. *FAM221A*, *IFIT5* and *TAS2R20* showed significant differential expression between HFpEF vs HFrEF patients in the right ventricle (**Figure 4**, **Supplementary Fig. 2**, **Supplementary Table 8**).

**Figure 4.**
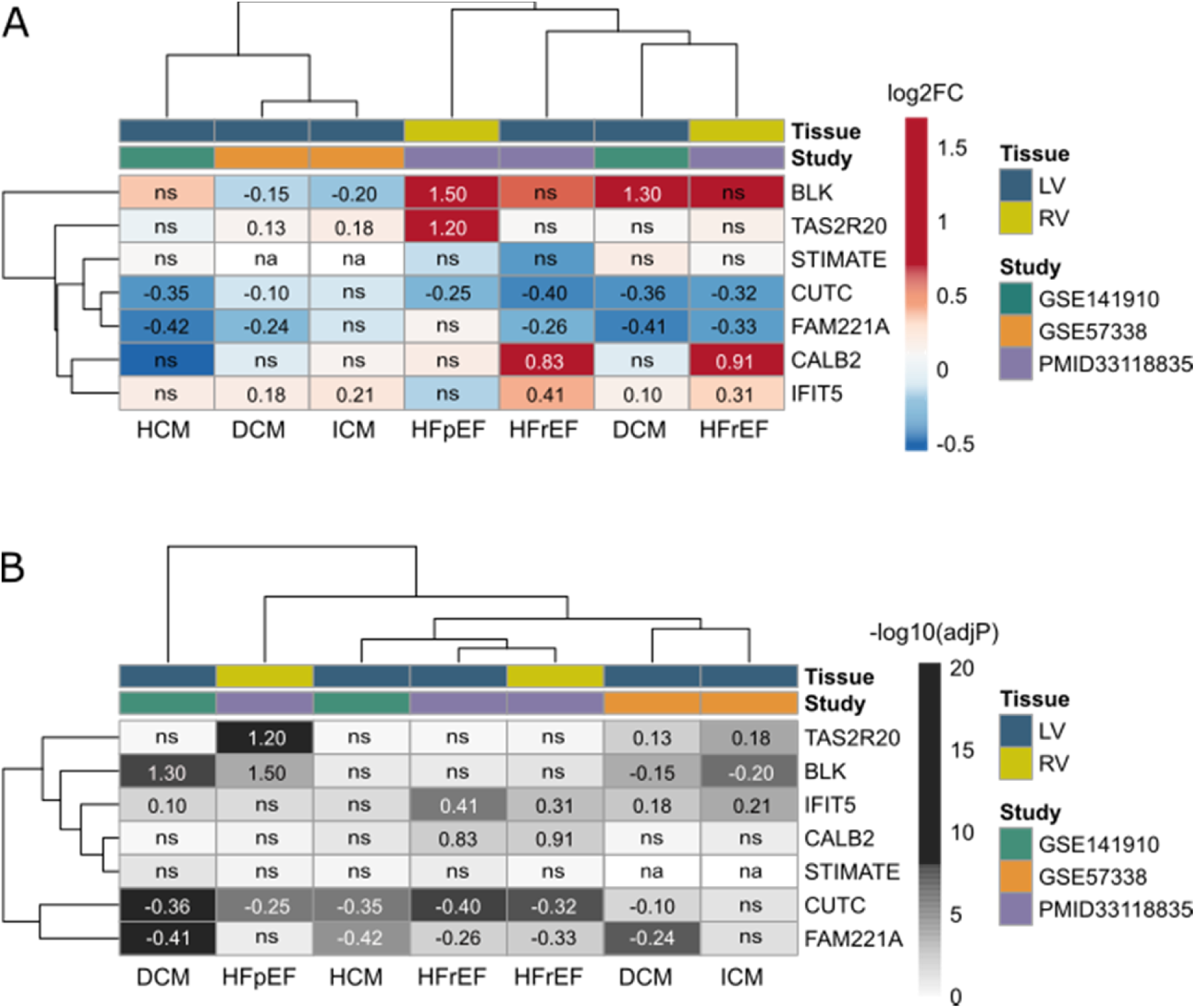
Heatmaps showing the differential expression in cardiac tissue from patients with heart failure vs controls of candidate genes from the meta-analysis of time to cardiovascular death and time to cardiovascular death and/or hospitalisation for heart failure. The heatmaps show **A**, magnitude of change (logFC; red: upregulated in HF vs control; blue: downregulated in HF vs control), and **B**, significance (-log_10_(adj. p-value)). ns: not significant (adj. p>0.05), na: not assessed.

Taken together, based on all transcriptional datasets, we highlight *CUTC* as a potential therapeutic target for heart failure progression that warrants further experimental validation and assessment.

### Functional enrichment analysis identifies molecular functions and pathways of candidate genes

To identify potential molecular functions and pathways underlying the genetic associations, we performed a gene ontology analysis of the candidate genes. We tested 1298 unique genes ranked by their lowest fixed model p-value across the ten non-synonymous collapsing analysis models from the time to cardiovascular death meta-analysis. We found enrichment (adjusted p<0.05) for the following molecular functions: adenyl ribonucleotide binding (GO:0032559, p=8.91×10^-4^); adenyl nucleotide binding (GO:0030554, p=1.43×10^-3^); lamin binding (GO:0005521, p=1.49×10^-3^); among others (**Supplementary Fig. 3A**, **Supplementary Table 9**).

We also performed the analysis for the 956 unique genes from the time to cardiovascular death and/or hospitalisation for heart failure meta-analysis. This analysis revealed enrichment for the following molecular functions and biological pathways: ion binding (GO:0043167, p=3.42×10^-3^); glycine, serine and threonine metabolism (KEGG:00260, p=4.17×10^-3^); among others (**Supplementary Fig. 3B**, **Supplementary Table 9**).

## Discussion

The identification of determinants of disease progression is critical for drug development, as most clinical trials assess the efficacy and safety of treatments that slow or arrest disease progression in patients with disease or prevent secondary events in specific patient groups. However, in contrast to the relatively large number of genetic studies on the incidence of common chronic diseases, including heart failure, there are few published studies investigating the genetic basis of disease progression.

In our study, we implemented a survival analysis for rare variants based on collapsing analysis and Cox proportional hazards regression with Firth’s penalized likelihood. We analysed the whole-exome sequencing data of 8,089 heart failure patients from three clinically well-defined studies: two clinical trials, CHARM and CORONA, and one population-based study: the UK Biobank. We performed meta-analysis and reported results found in at least two studies and supported by at least five carriers per study. We report seven genes not previously associated with heart failure progression with hazard ratios between 3.93 and 6.91. Specifically, we identified five candidate genes for cardiovascular death risk in heart failure patients: *FAM221A*, *CUTC*, *IFIT5*, *STIMATE* and *TAS2R20*, and two candidate genes for the composite of cardiovascular death and/or heart failure hospitalisation risk in heart failure patients: *CALB2* and *BLK*.

Importantly, none of these identified candidate gene loci have previously been associated with heart incidence through common-variant association studies, suggesting distinct genetic aetiology for heart failure progression compared to incidence. However, when evaluating the prior evidence of association with disease incidence, we found that all but two candidate gene loci (i.e., *FAM221A* and *TAS2R20*) harboured genome-wide significant, common-variant associations with cardiovascular traits, including heart rate, ascending aorta diameter and blood pressure.

We showed that several of the candidate genes are expressed in relevant cardiovascular tissues. In particular, *CUTC* is expressed strongly in heart tissues. Using heart single-cell RNA-seq data, we demonstrated that *CUTC* is expressed by all cardiac cell types with highest fractional expression in cardiomyocytes **(Supplementary Fig. 4**). Further, we demonstrated differential expression of five out of seven candidate genes between diseased and healthy hearts and/or between heart failure subtypes.

Finally, ML-based enrichment analysis supported our gene ranking from the meta-analysis, especially for the cardiovascular death outcome, as indicated by step-wise hypergeometric analyses. Functional enrichment analysis highlighted known pathways in cardiovascular disease, such as lamin binding (enrichment supported by *SUN1*, *TMEM201*, *SUN2*, *SYNE1* and *PLCB1*), and novel pathways, such as glycine, serine and threonine metabolism (supported by *GLDC*, *PGAM1* and *ALDH7A1*)^29, 30^.

*CUTC* was the only gene that showed an association with both tested endpoints, as well as strong expression in the heart and differential expression between diseased and healthy heart tissues. *CUTC* encodes a member of the CUT family of copper transporters that are associated with copper homeostasis and involved in the uptake, storage, delivery and efflux of copper^31–33^. Defective copper metabolism has been linked to various cardiovascular diseases, including heart failure. Mutations in genes encoding copper chaperones/transporters associate with cardiac disease in humans and mice^34^. Notably, the ongoing TRACER-HF trial (NCT03875183) evaluates the effects of a copper-binding agent INL1 in patients with HFrEF. INL1 is hypothesized to redistribute copper from high-concentration gradient (e.g., the circulation) to copper-depleted tissues (e.g., ischemic myocardial tissue), inducing heart regeneration. Although the exact function of CUTC remains to be determined, it has been suggested that it might act as an enzyme with Cu(I) as a cofactor rather than a copper transporter^31^. Moreover, CUTC participates in cardiac conduction and ion channel transport pathways based on the PathCards database (https://pathcards.genecards.org) and our co-expression network analysis suggested a role in muscle physiology and mitochondrial function (**Supplementary Fig. 4**). These pathways are all highly relevant biofunctions in heart failure pathophysiology. Our tractability assessment further revealed that CUTC contains a high-quality ligand pocket and is predicted druggable using small molecules (**Supplementary Table 10**). Taken together, *CUTC* may represent an attractive new candidate therapeutic target in heart failure.

Of note, alternative *CUTC* transcripts overlap *COX15*, encoding the Cytochrome C Oxidase Assembly Homolog. Despite a larger number of rare variant carriers than *CUTC*, *COX15* was not significantly associated with either cardiovascular death or the composite of cardiovascular death and/or heart failure hospitalisation in our analyses (**Supplementary Tables 11, 12**). Diseases associated with *COX15* include Mitochondrial Complex IV Deficiency, Nuclear Type 6 and Fatal Infantile Cardioencephalomyopathy Due To Cytochrome C Oxidase Deficiency. It has been proposed that *CUTC* and *COX15* functions in partnership as they share a bidirectional promoter^35^.

*STIMATE* and *CALB2* are involved in calcium binding, signalling or channel activity. Calcium pathways are fundamental for electrical signalling in the heart^36^. Several known mechanisms of cardiac pathologies are underlined by calcium channels^36^. A major class of drug targets already exist for calcium channel regulation^37, 38^. Calcium antagonists have been used to treat ischemia, a common cause of heart failure^39^. *IFIT5* and *BLK,* are involved in immune system function. It is established that inflammation plays an important role in chronic heart failure, after myocardial infarction or other myocardial damage, and heart failure is associated with circulating inflammatory cytokines that can predict clinical outcomes^40^. Whether inflammation plays a causal role remains to be established^41^. A number of taste receptors including *TAS2R20* are expressed in human heart and the role of these receptor in cardiac physiology and pathophysiology is not well understood yet^42^. It is notable that *TAS2R20* RNA expression is significantly increased in the myocardium in HFpEF patients but not in HFrEF patients, suggesting that *TAS2R20* may have a specific role in HFpEF.

In our meta-analysis, we did not discover any significantly associated protective genes. Such genes would be particularly promising for drug development as a protective effect of loss of function mimics the therapeutic inhibition of a drug. We observed previously in genetic analyses of heart failure incidence that detrimental association of rare variants are more prevalent than protective associations, because they are enriched for loss-of-function variants^14^.

We acknowledge that our study had limitations. Due to the paucity of exome-wide investigation of the role of rare variants in heart failure progression, we were currently unable to replicate our findings in an independent study cohort. More collaborative research is needed to address this limitation of our study, e.g., through the HERMES consortium that aims to assess the contribution of common genetic variants to heart failure progression^20^. Further, we note that the definition of heart failure amongst the cases was heterogeneous, for example, with respect to the presence or absence of cardiomyopathy, and different LVEF categories (HFrEF vs HFpEF). Due to the study of rare variants, stratifying patients according to phenotypic subtypes would have substantially reduced power. Additionally, for heart failure incidence and cardiomyopathy-associated rare variants, we previously detected some overlapping genetic architecture between heart failure subtypes^14^, supporting our combined approach in this study. Future larger studies would facilitate stratified analyses. Finally, our study focussed on individuals of European ancestry because there are insufficient numbers of patients of other ancestries. Further studies are needed to replicate our findings in other ancestry groups.

In conclusion, by rare-variant collapsing meta-analysis, we have identified seven candidate genes significantly associated with heart failure progression: *FAM221A, CUTC, IFIT5, STIMATE, TAS2R20, CALB2* and *BLK*. Pending replication, several of these genes could represent promising drug targets for heart failure, as supported by their tissue expression and function.

## Methods

### Ethics statement

Sites participating in the CHARM and CORONA studies received approval from local ethics committees for their conduct^22, 23^. Only patients who gave written informed consent for genetic analysis and for whom a DNA sample was available were included in the present study. The present study was performed in accordance with the policies on bioethics and human biologic samples of AstraZeneca. The protocols for UK Biobank are overseen by the UK Biobank Ethics Advisory Committee. For more information see https://www.ukbiobank.ac.uk/ethics/.

### CHARM and CORONA studies

The CHARM programme enrolled heart failure patients of >18 years of age into three distinct trials and randomly assigned them to receive candesartan or placebo: patients with LVEF >40% (CHARM Preserved; NCT00634712); patients with LVEF ≤ 40% and treated with angiotensin converting enzyme inhibitor (CHARM Added; NCT00634309); and patients with LVEF ≤ 40% and intolerant to angiotensin-converting enzyme inhibitor treatment (CHARM Alternative; NCT00634400)^22^. In the CHARM study, heart failure cause could be defined as ischemic, idiopathic, hypertensive or other causes^22^. The CORONA study enrolled patients of >60 years of age with chronic heart failure of ischemic cause and a LVEF ≤ 40% and randomly assigned them to receive rosuvastatin or placebo (NCT00206310)^23^. An overview of the patient characteristics of the trials is provided in **Table 1**.

In the CHARM and CORONA trials, the same two endpoints (i.e., cardiovascular death and a composite of cardiovascular death and/or heart failure hospitalisation) were defined by the clinical teams. For CHARM, cardiovascular death or unplanned admission to hospital for the management of worsening congestive heart failure was the primary outcome for the trial. All deaths were classified as cardiovascular unless an unequivocal non-cardiovascular cause was established. An heart failure hospital admission was defined as an admission to hospital because of heart failure with evidence of worsening heart failure^22^. For CORONA, the primary outcome was death from cardiovascular causes, nonfatal myocardial infarction, or nonfatal stroke. Deaths were classified as due to cardiovascular causes unless a definite non-cardiovascular reason was identified. Hospitalization for heart failure required documentation that worsening heart failure was the principal reason for hospitalization^23^.

### UK Biobank

The UK Biobank is a large prospective cohort study with 500,000 participants aged 40 to 69 years recruited from the general population in the United Kingdom^24^. The UK Biobank ICD10 codes (International Statistical Classification of Diseases and Related Health Problems, Tenth Revision codes) were interrogated for the following heart failure phenotypes: I11.0, I13.0, I13.2, I25.5, I42.0, I42.5, I42.8, I42.9 and I50.x^11^. Overall, 11,076 individuals with at least one primary or secondary hospital in-patient diagnoses code (Data-Field ID 41270), and/or at least one underlying (primary) cause of death in the death register (Data-Field ID 40001) in those categories, were considered as heart failure patients. In order to more closely match a clinical study design, only cases with heart failure diagnosis codes dated prior to recruitment were included in the analyses. Measurements of LVEF were not performed for the UK Biobank patients, as it was only available for a small number of individuals randomly selected from the biobank (**Table 1**).

For our analysis, we defined two endpoints: cardiovascular death (CVD) and a composite of cardiovascular death and/or heart failure hospitalisation (CVD_HFHOSP). In the UK Biobank, cardiovascular death was defined as a primary cause of death (Data-Field ID 40001) with any ICD10 codes in the Chapter IX (Diseases of the circulatory system), which correspond to I00-I99 codes. Heart failure hospitalisation was defined as a hospitalisation at least 28 days after the first diagnosis, with the same code used for the heart failure patient definition. To match the modelling approach used in the clinical studies, the date of recruitment was used to calculate the time to event. The end of follow-up was considered to be the date of the last major UK Biobank update (01/09/2019).

### Exome-sequencing

For CHARM, CORONA and UK Biobank, exomes were captured with the IDT xGen Exome Research Panel V1.0 (Integrated DNA Technologies, Coralville, IA, USA) and sequenced according to standard protocols on Illumina’s NovaSeq 6000 (Illumina, San Diego, CA, USA) platform. Exome sequencing for CHARM and CORONA was performed using 150bp paired-end reads at the Institute for Genomic Medicine at the Columbia University Medical Center, as previously described^14^. Exome sequencing for the UK Biobank using 75-bp paired-end reads and initial sample-level QC the UK Biobank was performed at Regeneron Pharmaceuticals, as previously described^14, 43, 44^. Quality control of the genetic data performed by Regeneron included sex discordance, contamination, unresolved duplicated sequences and discordance with microarray genotyping data^45^.

Sequence data from all three studies was processed through the AstraZeneca’s Centre for Genomics Research bioinformatics pipeline using a custom-built Amazon Web Services cloud compute platform running Illumina DRAGEN Bio-IT Platform Germline Pipeline v3.0.7. The reads were aligned to the GRCh38 genome reference, followed by single-nucleotide variant (SNV) and indel calling. SNVs and indels were first annotated using SnpEFF v4.34 against Ensembl Build 38.92. Variants were then annotated using the Genome Aggregation Database (gnomAD) minor allele frequencies (gnomAD v2.1.1 mapped to GRCh38), missense tolerance ratio (MTR) and REVEL scores^46–48^.

### Sample and variant quality control (QC)

The pre-QC dataset consisted of 3,090 CHARM, 2,906 CORONA and 2,868 UK Biobank participants. The following samples were removed: discordance between self-reported sex and genomic predicted sex based on X:Y coverage ratio; >4% contamination according to VerifyBamID v1.0.5; <95% of CCDS (release 22) bases covered with at least 10-fold coverage; related (up to third degree with KING v2.2.3), probability of European Ancestry <0.98 (<0.99 for UK Biobank) based on PEDDY v0.4.2 predictions; and individuals within four standard deviations of principal components 1-4 (**Supplementary Table 13**)^49–51^. The final filtered dataset for analysis consisted of 2,672 CHARM, 2,776 CORONA and 2,641 UK Biobank samples (**Figure 1**, **Table 1**).

### Gene-level collapsing analyses

We first performed, in the three cohorts separately, gene-based collapsing analyses to identify patients carrying at least one qualifying variant in each protein-coding gene. A qualifying variant is defined as a variant that passes certain filter criteria specific to each genetic model. **Supplementary Table 14** summarises the genetic models used for the analyses. We analysed ten non-synonymous models (nine of which are dominant and one recessive), plus an additional synonymous variant model as a negative control^52^.

The data was modelled using Cox proportional hazards regression with Firth’s penalized likelihood, which provides a solution to the non-convergence of the likelihood function, more frequently observed with rare-variant analysis: coxphf^53, 54^. Genotypes were encoded as binary variable after the first step of collapsing analysis. The coxphf method has been shown to be appropriate for genome-wide time-to-event analysis, both in terms of controlling type I error rates, and power for variants with low minor allele frequency^18^. In CHARM and CORONA, age at recruitment, sex and treatment arm were used as covariates in all analyses. The coxphf default of confidence intervals and tests based on the profile penalized log likelihood was used. In the UK Biobank, age at recruitment (Data-Field ID 21022) and sex (Data-Field 22001) were used as covariates in all analyses. For the collapsing survival analysis, the Bonferroni multiplicity adjusted threshold was set at p=2.7×10^-7^ (α = [0.05 / (18,500 genes * 10 non-synonymous models)]). This threshold was supported by permuting the data using R perm function (n=1) within the synonymous model, comparing with observed p-values with quantile-quantile plots and estimating the genomic inflation factor λ (**Supplementary Fig. 5**). The proportionality of hazards assumption was examined for the top genes by using the Schoenfeld residuals against the transformed time.

### Meta-analysis

We conducted a meta-analysis using PLINK v1.90 on the beta-values and standard errors extracted from the coxphf results. We computed both fixed and random effect meta-analysis models, and we only reported results supported by at least five carriers in each study. We computed both fixed and random effects meta-analysis models and reported random effects results with a heterogeneity index I^2^>40. Consistent with the survival analysis, the meta-analysis Bonferroni multiplicity adjusted threshold was set at p<2.7×10^-7^ (α = [0.05 / (18,500 genes * 10 non-synonymous models)]). Quantile-quantile plots for the meta-analysis were generated with (permutation-based) expected p-values from data permuted prior to the collapsing survival analysis using the R perm function (n=1) (**Supplementary Figs. 6A, 6B**).

### Sensitivity analysis

Analyses were conducted with additional covariates such as BMI and prior diabetes to assess the robustness of results when relevant for the genes with significant results (p<2.7×10^-7^) in the meta-analysis of CHARM, CORONA and the UK Biobank for both tested outcomes. Detailed clinical information supporting the analyses are reported in **Table 1**. Sensitivity analyses did not change the observed results (**Supplementary Table 15**).

### RNA expression

We interrogated median TPM-normalized gene expression for 54 tissues sampled in 948 donors provided by GTEx v8^27^. We defined a subset of 16 cardiovascular tissues as follows: Adipose - Subcutaneous, Adipose - Visceral (Omentum), Adrenal Gland, Artery - Aorta, Artery - Coronary, Artery - Tibial, Heart - Atrial Appendage, Heart - Left Ventricle, Kidney - Cortex, Kidney - Medulla, Liver, Muscle - Skeletal, Pancreas, Spleen, Thyroid, Whole Blood.

In addition, we extracted and analysed data from three published myocardial transcriptomic analyses: (1) HFpEF (n=41), HFrEF (n=30) and controls (n=24); (2) DCM (n=166), HCM (n=28) and controls (n=166) (GSE141910, The Myocardial Applied Genomics Network; https://www.med.upenn.edu/magnet/); and (3) idiopathic DCM (n=82), ICM (n=95) and controls (n=136) (GSE57338)^55^.

### Annotation with GWAS data

For the genes most significantly associated with heart failure progression, we compiled genetic evidence from the curated GWAS results of external studies available in the Cardiovascular Disease Knowledge Portal (https://cvd.hugeamp.org/). First, we listed the association with the lowest genome-wide significant p-values in the region, where associations are clumped by linkage disequilibrium and classified into three phenotypes group: cardiovascular, lipids and anthropometric. Then, we used the Cardiovascular Disease Knowledge Portal Genomic Region Miner tool to extract the lowest variant associations across each region or in the gene for the following six phenotypes: heart failure, non-ischemic cardiomyopathy, atrial fibrillation, any cardiovascular disease, myocardial infarction, or hypertension.

### Step-wise hypergeometric enrichment analysis with Mantis-ml

Mantis-ml is a method for gene prioritisation, leveraging publicly available gene annotation data from multiple resources to derive genetic association scores for a user-specified disease^25^. Harnessing data from resources such as OMIM, ExAC, Essential Mouse Genes, GnomAD, MSigDB, GTEx and genic-intolerance scores (RVIS and MTR) among others, we used this tool to assess to what extent the most significant genes from the meta-analysis are similar to genes known to be associated with heart failure or cardiomyopathy.

We performed a step-wise hypergeometric test between the study-ranked list of genes with a fixed p-value<0.01 for one of the outcomes (**Supplementary Tables 11, 12**) and 18,626 genes pre-ranked by their Mantis-ml association scores for 33 diseases containing either ‘heart failure’ or ‘cardiomyopathy’ (i.e., 26 from OpenTargets, four from HPO, three from Genomics England). An overview of these diseases is provided in **Supplementary Table 16**. The resulting step-wise hypergeometric curves are shown in **Supplementary Fig. 1**.

This test quantifies the overlap between the list of genes identify in this heart failure progression study and the list of genes ranked according to independent data sources by Mantis-ml. The p-values generated are converted to Phred scores. Enrichment performance were assessed and p-values obtained using the one-sided Mann-Whitney U test.

### Gene ontology enrichment analysis

A functional enrichment analysis was performed using g:Profiler for the same lists of genes ordered on fixed p-value as done for the annotation with Mantis-ml, i.e., the genes with a fixed effect p-value<0.01 for each of the outcomes (**Supplementary Tables 11, 12**)^56^. The data sources tested were Gene Ontology (GO, molecular function, cellular component, biological process), KEGG and miRTarBase. The background set of gene used to compute the functional enrichment was the 18,948 genes list from the Consensus Coding Sequence (CCDS, release 22), over annotated genes. Multiple testing correction for p-values was computed by applying the default g:SCS method.

### Tractability assessment

For the genes most significantly associated with heart failure progression, we assessed the amenability to intervention by different drug modalities, including small molecules, antibodies, PROTAC and others, using the Open Target Platform (https://platform.opentargets.org/). The genes were categorized based on ‘buckets’ representing different levels of tractability, ranging from high confidence (lower number) to uncertain tractability (higher number), as previously described^57^. In addition, genes were also classified according to their ‘target development level’ (TDL) into either of four classes (i.e., Tclin, Tchem, Tbio or Tdark) using the Target Central Resource Database (TCRD, http://juniper.health.unm.edu/tcrd/). In brief, Tclin are proteins through which approved drugs act (i.e., mode-of-action drug targets); Tchem are proteins known to bind small molecules with high potency; Tbio are proteins with well-studied biology, having a fractional publication count above 5; and Tdark are understudied proteins that do not meet criteria for the above 3 categories, respectively (**Supplementary Table 10**).

### Co-expression network analysis

To further explore the *CUTC* function, we performed co-expression analysis using GeneNetwork v2.0 (https://www.genenetwork.nl/), which predicts pathway and human phenotype associations using 31,499 public human RNA-seq samples (**Supplementary Fig. 4**).^58^

## Data availability

Summary statistics from the rare-variant collapsing analyses are available within the article and its Supplementary Information files. Access to the UK Biobank data can be gained via the UK Biobank website: https://bbams.ndph.ox.ac.uk/ams/.

## Code availability

Sequence data were processed through a custom-built Amazon Web Services cloud compute platform running Illumina DRAGEN Bio-IT Platform. SNVs and indels were annotated using SnpEFF v4.3 against Ensembl v38.92. The software used in this study are referenced in the manuscript and are available online: Mantis-ml (https://github.com/astrazeneca-cgr-publications/mantis-ml-release), QQperm (https://cran.r-project.org/web/packages/QQperm/index.html).

## Acknowledgments

We thank the participants and investigators of the UK Biobank study who made this work possible (Resource Application Number 26041). We thank the UKB Exome Sequencing Consortium (UKB-ESC) members AbbVie, Alnylam Pharmaceuticals, AstraZeneca, Biogen, Bristol-Myers Squibb, Pfizer, Regeneron and Takeda for funding the generation of the data, and the Regeneron Genetics Center for completing the sequencing and initial quality control of the exome sequencing data. We acknowledge the AstraZeneca Centre for Genomics Research Analytics and Informatics team for processing and analysis of sequencing data. We thank all the study participants for contributing to this effort and the CHARM and CORONA co-investigators. Finally, we thank Sri Deevi and Dorota Matelska from the AstraZeneca Centre for Genomics Research for their support, for which we are grateful. M.-P.D. is funded by the Canada Research Chairs Program and by the Health Collaboration Acceleration Fund from the Government of Quebec. J.C.T. holds the Canada Research Chair in personalized and translational medicine and the Université de Montréal endowed research chair in atherosclerosis.

## Author Contributions Statement

O.C., M-P.D., C.H., J-C.T., D.S.P. and K.C. conceived and designed the study. O.C., Q.W., L.M. and A.W. performed the statistical and computational analyses. M-P.D., D.V., D.S.P. and K.C. supervised the analyses. M-P.D., C.B.G, J.K., C.H., J-C.T., D.S.P. and K.C. participated in the acquisition of the data. O.C., M-P.D., Q.W., L.M., D.V., A.W., Q-D.W., K.M.H., C.B.G, J.K., C.H., J-C.T., D.S.P. and K.C. participated in the data interpretation. O.C., M-P.D., L.M., A.W., J-C.T., D.S.P. and K.C. wrote the initial draft of the manuscript. All authors contributed to the revision of the first draft. All authors approved the final version of the manuscript.

## References

1 GBD 2017 Disease and Injury Incidence and Prevalence Collaborators. Global, regional, and national incidence, prevalence, and years lived with disability for 354 diseases and injuries for 195 countries and territories, 1990-2017: a systematic analysis for the Global Burden of Disease Study 2017. Lancet 392, 1789–1858 (2018).

2 Metra, M. & Teerlink, J. R. Heart failure. Lancet 390, 1981–1995 (2017).

3 McMurray, J. J. V. & Pfeffer, M. A. Heart failure. Lancet 365, 1877–1889 (2005).

4 Dickstein, K. et al. ESC Guidelines for the diagnosis and treatment of acute and chronic heart failure 2008: the Task Force for the Diagnosis and Treatment of Acute and Chronic Heart Failure 2008 of the European Society of Cardiology. Developed in collaboration with the Heart Failure Association of the ESC (HFA) and endorsed by the European Society of Intensive Care Medicine (ESICM). Eur Heart J 29, 2388–2442 (2008).

5 Cook, C., Cole, G., Asaria, P., Jabbour, R. & Francis, D. P. The annual global economic burden of heart failure. Int J Cardiol 171, 368–376 (2014).

6 Savarese, G. & Lund, L. H. Global Public Health Burden of Heart Failure. Card Fail Rev 3, 7–11 (2017).

7 Retrum, J. H. et al. Patient-identified factors related to heart failure readmissions. Circ Cardiovasc Qual Outcomes 6, 171–177 (2013).

8 Roger, V. L., et al. Heart disease and stroke statistics--2012 update: a report from the American Heart Association. Circulation 125, e2–e220 (2012).

9 National Clinical Guideline Centre (UK). Chronic Heart Failure: National Clinical Guideline for Diagnosis and Management in Primary and Secondary Care: Partial Update. (Royal College of Physicians (UK), 2010).

10 Lindgren, M. P. et al. A Swedish Nationwide Adoption Study of the Heritability of Heart Failure. JAMA cardiology 3, 703–710 (2018).

11 Shah, S. et al. Genome-wide association and Mendelian randomisation analysis provide insights into the pathogenesis of heart failure. Nature Communications 11, 163 (2020).

12 Arvanitis, M. et al. Genome-wide association and multi-omic analyses reveal ACTN2 as a gene linked to heart failure. Nature Communications 11, 1122 (2020).

13 Joseph, J. et al. Genetic Architecture of Heart Failure with Preserved versus Reduced Ejection Fraction. medRxiv, 2021.2012.2001.21266829 (2021).

14 Povysil, G. et al. Assessing the Role of Rare Genetic Variation in Patients With Heart Failure. JAMA Cardiol 6, 379–386 (2021).

15 Cox, D. R. Regression Models and Life-Tables. Journal of the Royal Statistical Society: Series B (Methodological) 34, 187–202 (1972).

16 Xue, X. et al. Testing the proportional hazards assumption in case-cohort analysis. BMC Med Res Methodol 13, 88 (2013).

17 Li, R. et al. Fast Lasso method for large-scale and ultrahigh-dimensional Cox model with applications to UK Biobank. Biostatistics 23, 522–540 (2022).

18 Bi, W., Fritsche, L. G., Mukherjee, B., Kim, S. & Lee, S. A Fast and Accurate Method for Genome-Wide Time-to-Event Data Analysis and Its Application to UK Biobank. American Journal of Human Genetics 107, 222–233 (2020).

19 Legault, M.-A., Perreault, L.-P. L. & Dubé, M.-P. ExPheWas: a browser for gene-based pheWAS associations. medRxiv, 2021.2003.2017.21253824 (2021).

20 Lumbers, R. T. et al. The genomics of heart failure: design and rationale of the HERMES consortium. ESC Heart Fail 8, 5531–5541 (2021).

21 Povysil, G. et al. Rare-variant collapsing analyses for complex traits: guidelines and applications. Nat Rev Genet 20, 747–759 (2019).

22 Pfeffer, M. A. et al. Effects of candesartan on mortality and morbidity in patients with chronic heart failure: the CHARM-Overall programme. Lancet 362, 759–766 (2003).

23 Kjekshus, J. et al. Rosuvastatin in older patients with systolic heart failure. N Engl J Med 357, 2248–2261 (2007).

24 Bycroft, C. et al. The UK Biobank resource with deep phenotyping and genomic data. Nature 562, 203–209 (2018).

25 Vitsios, D. & Petrovski, S. Mantis-ml: Disease-Agnostic Gene Prioritization from High-Throughput Genomic Screens by Stochastic Semi-supervised Learning. American Journal of Human Genetics 106, 659–678 (2020).

26 Tobita, T. et al. Genetic basis of cardiomyopathy and the genotypes involved in prognosis and left ventricular reverse remodeling. Sci Rep 8, 1998 (2018).

27 Consortium, G. T. The GTEx Consortium atlas of genetic regulatory effects across human tissues. Science 369, 1318–1330 (2020).

28 Hahn, V. S. et al. Myocardial Gene Expression Signatures in Human Heart Failure With Preserved Ejection Fraction. Circulation 143, 120–134 (2021).

29 Worman, H. J., Ostlund, C. & Wang, Y. Diseases of the nuclear envelope. Cold Spring Harb Perspect Biol 2, a000760 (2010).

30 Wittemans, L. B. L. et al. Assessing the causal association of glycine with risk of cardio-metabolic diseases. Nature Communications 10, 1060 (2019).

31 Li, Y., Du, J., Zhang, P. & Ding, J. Crystal structure of human copper homeostasis protein CutC reveals a potential copper-binding site. J Struct Biol 169, 399–405 (2010).

32 Gupta, S. D., Lee, B. T., Camakaris, J. & Wu, H. C. Identification of cutC and cutF (nlpE) genes involved in copper tolerance in Escherichia coli. J Bacteriol 177, 4207–4215 (1995).

33 Li, J. et al. Identification and characterization of a novel Cut family cDNA that encodes human copper transporter protein CutC. Biochem Biophys Res Commun 337, 179–183 (2005).

34 Liu, Y. & Miao, J. An Emerging Role of Defective Copper Metabolism in Heart Disease. Nutrients 14 (2022).

35 Uchiumi, F., Fujikawa, M., Miyazaki, S. & Tanuma, S.-i. Implication of bidirectional promoters containing duplicated GGAA motifs of mitochondrial function-associated genes. AIMS Mol Sci 1, 1–26 (2014).

36 Berridge, M. J., Bootman, M. D. & Roderick, H. L. Calcium signalling: dynamics, homeostasis and remodelling. Nat Rev Mol Cell Biol 4, 517–529 (2003).

37 Rinnier, R. T., Goldberg, M. E. & Torjman, M. C. in The Essence of Analgesia and Analgesics (eds J. Michael Watkins-Pitchford, Jonathan S. Jahr, & Raymond S. Sinatra) 310–315 (Cambridge University Press, 2010).

38 Suckfüll, M. et al. A randomized, double-blind, placebo-controlled clinical trial to evaluate the efficacy and safety of neramexane in patients with moderate to severe subjective tinnitus. BMC Ear Nose Throat Disord 11, 1 (2011).

39 Sueta, D., Tabata, N. & Hokimoto, S. Clinical roles of calcium channel blockers in ischemic heart diseases. Hypertens Res 40, 423–428 (2017).

40 Yndestad, A. et al. Role of inflammation in the progression of heart failure. Curr Cardiol Rep 9, 236–241 (2007).

41 Dick, S. A. & Epelman, S. Chronic Heart Failure and Inflammation: What Do We Really Know? Circ Res 119, 159–176 (2016).

42 Bloxham, C. J., Foster, S. R. & Thomas, W. G. A Bitter Taste in Your Heart. Front Physiol 11, 431 (2020).

43 Wang, Q. et al. Rare variant contribution to human disease in 281,104 UK Biobank exomes. Nature 597, 527–532 (2021).

44 Szustakowski, J. D. et al. Advancing human genetics research and drug discovery through exome sequencing of the UK Biobank. Nat Genet 53, 942–948 (2021).

45 Van Hout, C. V. et al. Exome sequencing and characterization of 49,960 individuals in the UK Biobank. Nature 586, 749–756 (2020).

46 Ioannidis, N. M. et al. REVEL: An Ensemble Method for Predicting the Pathogenicity of Rare Missense Variants. Am J Hum Genet 99, 877–885 (2016).

47 Traynelis, J. et al. Optimizing genomic medicine in epilepsy through a gene-customized approach to missense variant interpretation. Genome Res 27, 1715–1729 (2017).

48 Karczewski, K. J. et al. The mutational constraint spectrum quantified from variation in 141,456 humans. Nature 581, 434–443 (2020).

49 Pedersen, B. S. & Quinlan, A. R. Who’s Who? Detecting and Resolving Sample Anomalies in Human DNA Sequencing Studies with Peddy. American Journal of Human Genetics 100, 406–413 (2017).

50 Jun, G. et al. Detecting and estimating contamination of human DNA samples in sequencing and array-based genotype data. American Journal of Human Genetics 91, 839–848 (2012).

51 Manichaikul, A. et al. Robust relationship inference in genome-wide association studies. *Bioinformatics (Oxford*, England) 26, 2867–2873 (2010).

52 Carss, K. J. et al. Spontaneous Coronary Artery Dissection: Insights on Rare Genetic Variation From Genome Sequencing. Circ Genom Precis Med 13, e003030 (2020).

53 Heinze, G. & Schemper, M. A solution to the problem of monotone likelihood in Cox regression. Biometrics 57, 114–119 (2001).

54 Dunkler, D., Ploner, M., Schemper, M. & Heinze, G. Weighted Cox Regression Using the R Package coxphw. J. Stat. Soft. 84, 1–26 (2018).

55 Liu, Y. et al. RNA-Seq identifies novel myocardial gene expression signatures of heart failure. Genomics 105, 83–89 (2015).

56 Raudvere, U. et al. g:Profiler: a web server for functional enrichment analysis and conversions of gene lists (2019 update). Nucleic Acids Research 47, W191–W198 (2019).

57 Brown, K. K. et al. Approaches to target tractability assessment - a practical perspective. Medchemcomm 9, 606–613 (2018).

58 Deelen, P. et al. Improving the diagnostic yield of exome-sequencing by predicting gene-phenotype associations using large-scale gene expression analysis. Nat Commun 10, 2837 (2019).

59 Cochran, W. G. The Combination of Estimates from Different Experiments. Biometrics 10, 101–129 (1954).

60 Higgins, J. P. T. & Thompson, S. G. Quantifying heterogeneity in a meta-analysis. Stat Med 21, 1539–1558 (2002).

61 Cerami, E. et al. The cBio cancer genomics portal: an open platform for exploring multidimensional cancer genomics data. Cancer Discov 2, 401–404 (2012).

62 Gao, J. et al. Integrative analysis of complex cancer genomics and clinical profiles using the cBioPortal. Sci Signal 6, pl1 (2013).

